# ARTIFICIAL INTELLIGENCE ACCURATELY DETECTS TRAUMATIC THORACOLUMBAR FRACTURES ON SAGITTAL RADIOGRAPHS

**DOI:** 10.1101/2021.05.09.21256762

**Authors:** Guillermo Sanchez Rosenberg, Andrea Cina, Giuseppe Rosario Schirò, Pietro Domenico Giorgi, Boyko Gueorguiev, Mauro Alini, Peter Varga, Fabio Galbusera, Enrico Gallazzi

## Abstract

**Background context:** Traumatic thoracolumbar (TL) fractures are frequently encountered in emergency rooms. Sagittal and anteroposterior radiographs are the first step in the trauma routine imaging. Up to 30% of TL fractures are missed in this imaging modality, thus requiring a CT and/or MRI to confirm the diagnosis. A delay in treatment leads to increased morbidity, mortality, exposure to ionizing radiation and financial burden. Fracture detection with Machine Learning models has achieved expert level performance in previous studies. Reliably detecting vertebral fractures in simple radiographic projections would have a significant clinical and financial impact.

**Purpose:** To develop a deep learning model that detects traumatic fractures on sagittal radiographs of the TL spine.

**Study design/setting:** Retrospective Cohort study.

**Methods:** We collected sagittal radiographs, CT and MRI scans of the TL spine of 362 patients exhibiting traumatic vertebral fractures. Cases were excluded when CT and/or MRI where not available. The reference standard was set by an expert group of three spine surgeons who conjointly annotated the sagittal radiographs of 171 cases. CT and/or MRI were reviewed to confirm the presence and type of the fracture in all cases. 302 cropped vertebral images were labelled ‘fracture’ and 328 ‘no fracture’. After augmentation, this dataset was then used to train, validate, and test deep learning classifiers based on ResNet18 and VGG16 architectures. To ensure that the model’s prediction was based on the correct identification of the fracture zone, an Activation Map analysis was conducted.

**Results:** Vertebras T12 to L2 were the most frequently involved, accounting for 48% of the fractures. A4, A3 and A1 were the most frequent AO Spine fracture types. Accuracies of 88% and 84% were obtained with ResNet18 and VGG16 respectively. The sensitivity was 89% with both architectures but ResNet18 showed a higher specificity (88%) compared to VGG16 (79%). The fracture zone was precisely identified in 81% of the heatmaps.

**Conclusions:** Our AI model can accurately identify anomalies suggestive of vertebral fractures in sagittal radiographs by precisely identifying the fracture zone within the vertebral body.

**Clinical significance:** Clinical implementation of a diagnosis aid tool specifically trained for TL fracture identification is anticipated to reduce the rate of missed vertebral fractures in emergency rooms.

## Introduction

The thoracolumbar (TL) spine is one of the most common site of traumatic fracture occurrence, with an incidence that ranges from 32 to 64/100.000 per year; furthermore, traumatic TL fractures have a rate of associated neurological injuries from 22% to 51% depending on the fracture type, and a require surgical treatment in 38% of the cases. (1–3) Traumatic TL fractures are associated with decreased physical function, severe reduction of quality of life and the lowest rate of return to work among all major organ injuries. (4) Additionally, the overall mortality associated to spinal injuries is 17%. (5) Besides this elevated disease burden and prevalence, the treatment of vertebral fractures is costly. The annual estimated economic cost, in the United States alone, surpassed the billion dollar figure already in 2011. (6)

The severity of traumatic TL fractures can range from a simple apophyseal fracture without structural impairment to a complete dislocation of the spine. Sagittal and anteroposterior radiographs are the first step in the trauma routine imaging. However, they are not a very reliable diagnosis aid when suspecting TL fractures: the worldwide reported false-negative rate is as high as 30%. (7) Moreover, the current classification systems such as the AOSpine Classification and the Thoracolumbar Injury Classification and Severity score (TLICS) stratify fracture severity on parameters such as anterior failure of the vertebral disc under compression and posterior integrity of ligamentous structures (8), which are not readily identifiable in radiographs. The severity and instability of the fracture will determine the need for conservative or surgical treatment. Thus, surgeons must resort to second level imaging such as Computer Tomography (CT) or Magnetic Resonance Imaging (MRI) to determine the treatment strategy. (9) The need for second level imaging inevitably leads to a delay in diagnosis, ranging from hours to even months, frequently resulting in poorer clinical outcomes. (10,11)

The recent explosion of labeled data, namely ‘big data’, has brought upon the era of artificial intelligence (AI). Within the healthcare sector, AI is being now used for several applications including drug discovery, remote patient monitoring, risk management, wearables, virtual assistants, and hospital management. Regarding medical diagnostics and imaging, the field of radiology has been particularly benefited. (12) The management of patients with musculoskeletal diseases could be improved by these innovations, provided that optimal accuracies are preserved. (13–15) By supporting the treating physician in identifying anomalies on patients imaging studies, AI is posed to considerably reduce diagnostic errors (16), therefore improving clinical outcomes in the treatment of vertebral fractures.

Deep learning (DL) is a supervised machine learning method that uses an algorithmic structure based on neural networks, such as Convolutional Neural Networks (CNN) (17). This method has been reported to perform as good or even better than humans in image classification (18). The power of this technique lies int the ability to identify and extract relevant features from labeled data at a grand scale. (19)

Recently, several proof of concept papers were published showing the application of AI and DL in spine imaging, showing promising results in the evaluation of degenerative disorders (20), adult deformities (21), adolescent idiopathic scoliosis (22), detection of primary and secondary bone tumors (23,24), and vertebral fractures (25).

Implementing a diagnosis aid tool specially trained for fracture identification in the clinical practice is anticipated to reduce the rate of missed vertebral fractures in emergency rooms. Murata et al have recently published a model capable of detecting vertebral fractures in plain radiographs. (26) However, Murata’s study is limited to radiographs displaying only one vertebral fracture, therefore invalidating its application in a scenario where multiple are present. Of note, the annotation of each image subgroup was done by a single spine surgeon, which could result in the introduction of confirmation bias in the standard of reference.

The main aim of this study was to develop an AI-based algorithm capable of accurately detecting vertebral fractures in sagittal radiographs of the TL spine. The secondary aim was to gain a deeper understanding of the model’s interpretation of the ‘fracture zone’ through a heatmap representation.

## Methods

This work utilized a CNN-based supervised learning approach.(17) The model was trained on cropped single vertebrae. The standard of reference was set by three expert spine surgeons, who evaluated the plain sagittal radiograph together with second level imaging, namely CT and/or MRI and annotated the single vertebra image as ‘fracture’ or ‘non-fracture’.

### Image acquisition and standard of reference

Sagittal radiographs, CT and MRI scans from the TL spine of 362 patients of more than 12 years of age and exhibiting traumatic vertebral fractures were retrospectively collected from a Spine Surgery reference Center (ASST Grande Ospedale Metropolitano Niguarda, Milano, Italy). The Ethical Committee approval was granted via the *Comitato Etico Milano Area 3* under the Ref. No. 359-24062020. To ensure accuracy of the diagnoses, only cases with an initial radiograph and a subsequent CT or MRI were included. Fractures resulting from mechanisms other than trauma, such as osteoporosis or pathologic fractures were excluded. Sagittal radiographs and second level imaging of 151 patients were selected for the final analysis. An expert group of three spine surgeons with more than 30 years of accumulated experience reviewed each case individually to identify the presence of a fracture, and then classified them according to the AO Spine Classification. Finally, in the cases where disagreement existed, meetings were held to reach unanimous consensus for each case.

A total of 630 single vertebra images, obtained from in 222 sagittal radiographs of the TL spine were annotated. Of these, 302 were annotated as ‘fracture’ and 328 as ‘non-fracture’. The annotation process was performed using a C++ software specifically developed for this project. The annotator indicated the Region of Interest (ROI) with a bounding box around a vertebra, and assigned the class, ‘fracture’ or ‘non-fracture’, the level and the corresponding AO Spine Classification.

### Training and Test Sets

The image dataset was split into a training set (N = 578) and a test set (N = 52). To increase the generalization capability of the model, we used augmentation techniques such as random rotation, flipping, and shifting. In this way the model was trained on different versions of the same image during the training epochs.

### Classification with Deep Learning

We compared the performances of VGG16 (27) and ResNet18 (28) DL architectures, which were in the top three of the ImageNet challenge (29) in 2014 and 2015 respectively. These CNNs achieved state-of-the-art performances on computer vision tasks such as image classification, object detection, and landmark localization. (25) The difference between these CNNs is that VGG16 is a plain neural network with a deep sequence of convolutional layers followed by max-pooling, while ResNet functions with the so-called residual blocks, where the input of each block is summed to the output of the same block creating a skip connection **(Fig. 1)**. The motivation behind the skipping connection is the vanishing gradient problem that arises during the training of very deep CNNs. (27) The images were resized to 512×512 pixels and normalized to have zero mean and unit variance, according to the image guidelines used in the ImageNet challenge.

**Figure 1:**
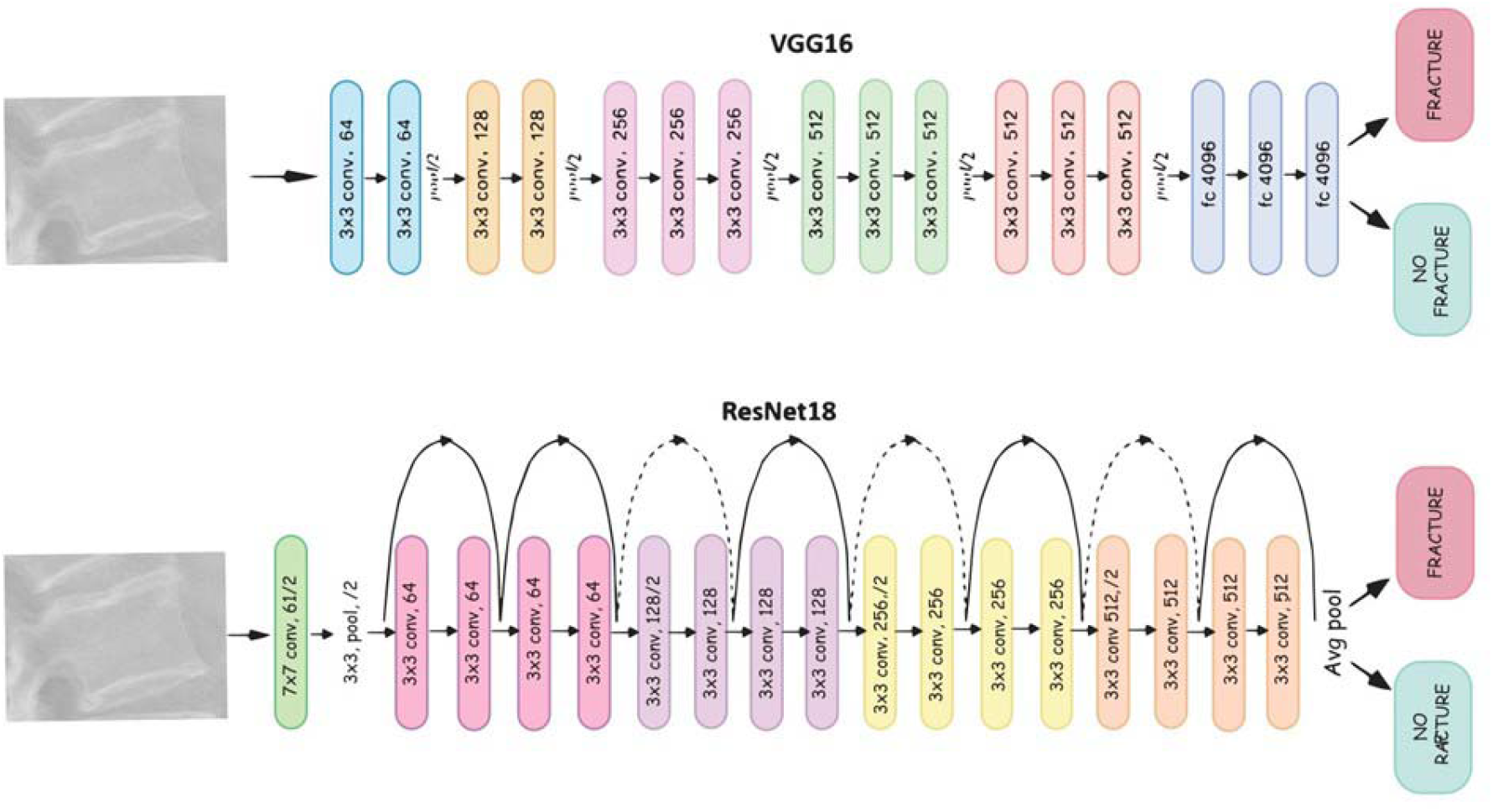
DL network architectures used in this study. The VGG16 is a sequence of convolution and max-pooling operations. The number of parameters to learn is remarkably high (about 138 million). ResNet18 presents the so-called Residual Blocks that represent the blocks of convolution operation between the skipping connections represented by the arrows, reducing the number of parameters to approximately 11 million.

Since the number of available images was not high, we used a technique called Transfer Learning (30,31) that consists of exploiting parameters of the 2 models that obtained state-of-the-art performances on the ImageNet challenge and retrained only the last few layers, the 2 last residual blocks in the present study, on the new task of vertebral fracture classification. We replaced the last fully connected layers of both networks (with 1000 neurons each) with a fully connected layer with 2 neurons representing the 2 classes. For the fully connected layer we used the softmax activation function that returns a probability distribution that assigns to each sample a probability of belonging to a class. The objective function is the negative log-likelihood loss used together with the softmax.

To find the best hyperparameters to train the network, in particular the learning rate and the batch size, we performed cross-validation with 10 folds where the training set is split into 10 parts, 9 of which are used to train the model and 1 for validation. This process was repeated 10 times iteratively until all the combinations of the folds have been used for the training/validation process. Finally, the entire training set was used for training with the best hyperparameters found in the cross-validation and we evaluated the performances of the model on the test set.

The model was implemented in Python language using PyTorch (34), a deep learning framework developed by Facebook. For the training and the evaluation, we used a Linux workstation with a NVIDIA QUADRO RTX 5000. The models ran for 200 epochs using a batch size of 32 and a learning rate of 0.00016, which resulted as the best hyperparameters in the cross-validation step. We used the Adam optimizer for model optimization and a method that reduced the learning rate by a factor of 0.1 if the accuracy did not improve for 10 epochs in a row (ReduceLROnPlateau in PyTorch).

### Evaluation

#### Model’s Performance Parameters

The models’ performance was assessed quantitatively by calculating the accuracy, sensitivity, and specificity in fracture identification. Accuracy represents the ability of the model to assign the images to the correct class, described by the AUC **(Fig. 5)**; the AUC outputs a value that displays the probability of a random sample being correctly classified by the algorithm, thus indicating the capacity of a classifier to distinguish between two classes. (32) Sensitivity describes the ability to detect the fractures, and specificity is the ability to detect the lack of a fracture.

#### Understanding the Model’s Prediction

To ensure that the model’s prediction was based on the correct identification of the fracture zone, we implemented Activation Maps on the single vertebral images to highlight the image regions that lead the model to classify an image into a specific class. The activation maps were obtained by multiplying the second last layer of the neural network (the last feature maps) by the weights that point to the neuron of the class predicted by the model. This way, all pixels of the feature maps were weighted according to the model prediction highlighting the most important parts of the images that determined the prediction. The heatmaps were evaluated by the same surgeons that set the standard of reference, judging whether the “warm zones” seen in the Activation Maps correlated to the fracture zones seen the CT or MRI images.

## Results

### Clinical Dataset

For the final analysis, a total of 151 cases of patients with TL fracture with availability of the initial radiograph and subsequent CT or MRI were selected. A total of 222 TL sagittal radiographs were analyzed and classified by the expert group of spine surgeons. Vertebras T12 to L2 were the most frequently involved, accounting for 48% of the fractures **(Fig. 2)**. Axial compression fractures, namely the AO Spine types A4, A3 and A1, were the most frequent injury mechanisms **(Fig. 3)**.

**Figure 2:**
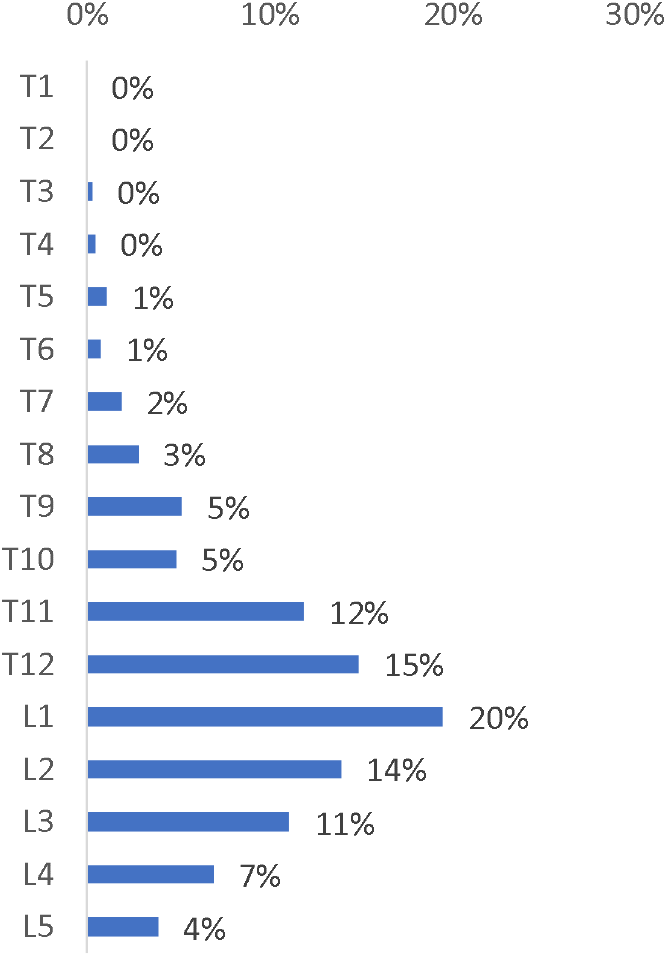
Epidemiological distribution of TL fractures.

**Figure 3:**
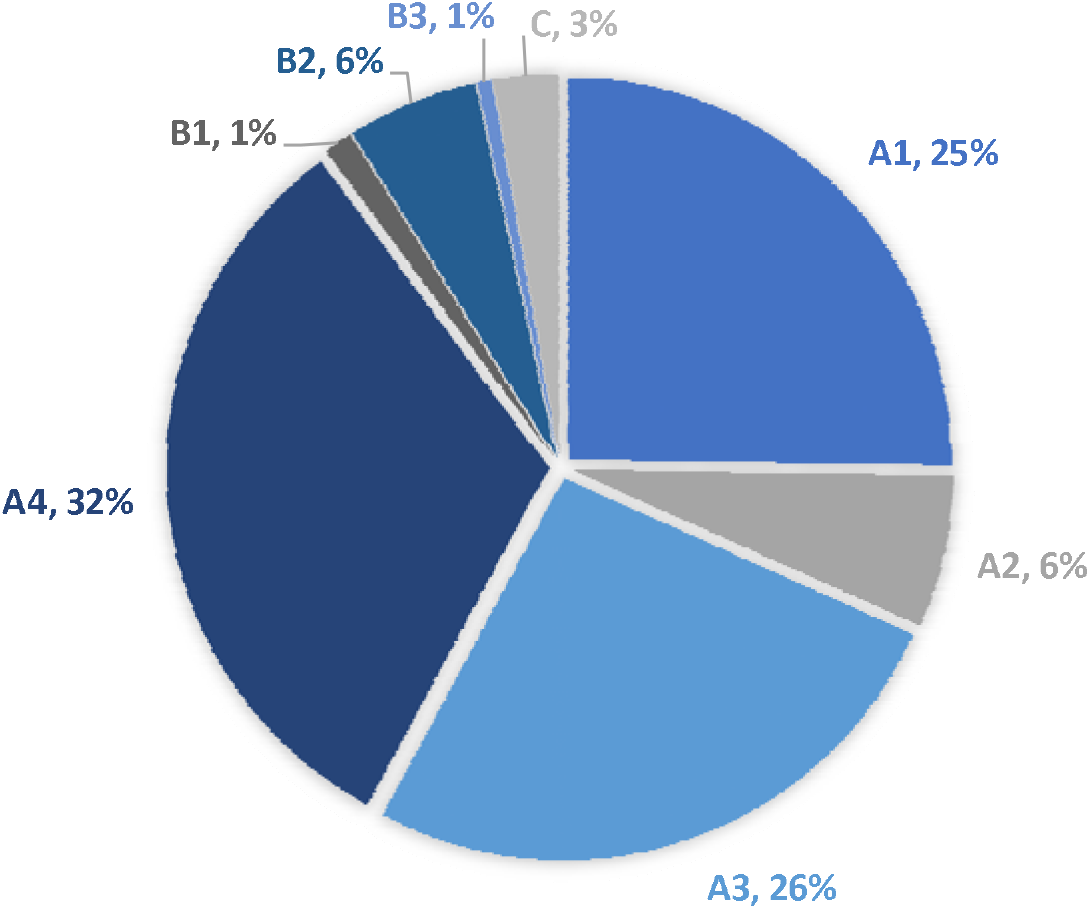
TL fracture type per AO Classification.

### Deep Learning Model

Both DL architectures achieved high accuracy, sensitivity, and specificity after hyperparameter optimization, but ResNet18 performed better in all these aspects compared to VGG16. Both models predicted three false negatives (5.8%) by misclassifying three ‘fracture’ images as ‘no fracture’. ResNet18 showed increased specificity, predicting three false positives in comparison to the 5 of the VGG16, namely classifying ‘no fracture’ images as ‘fracture’. **(Fig. 4)**.

**Figure 4:**
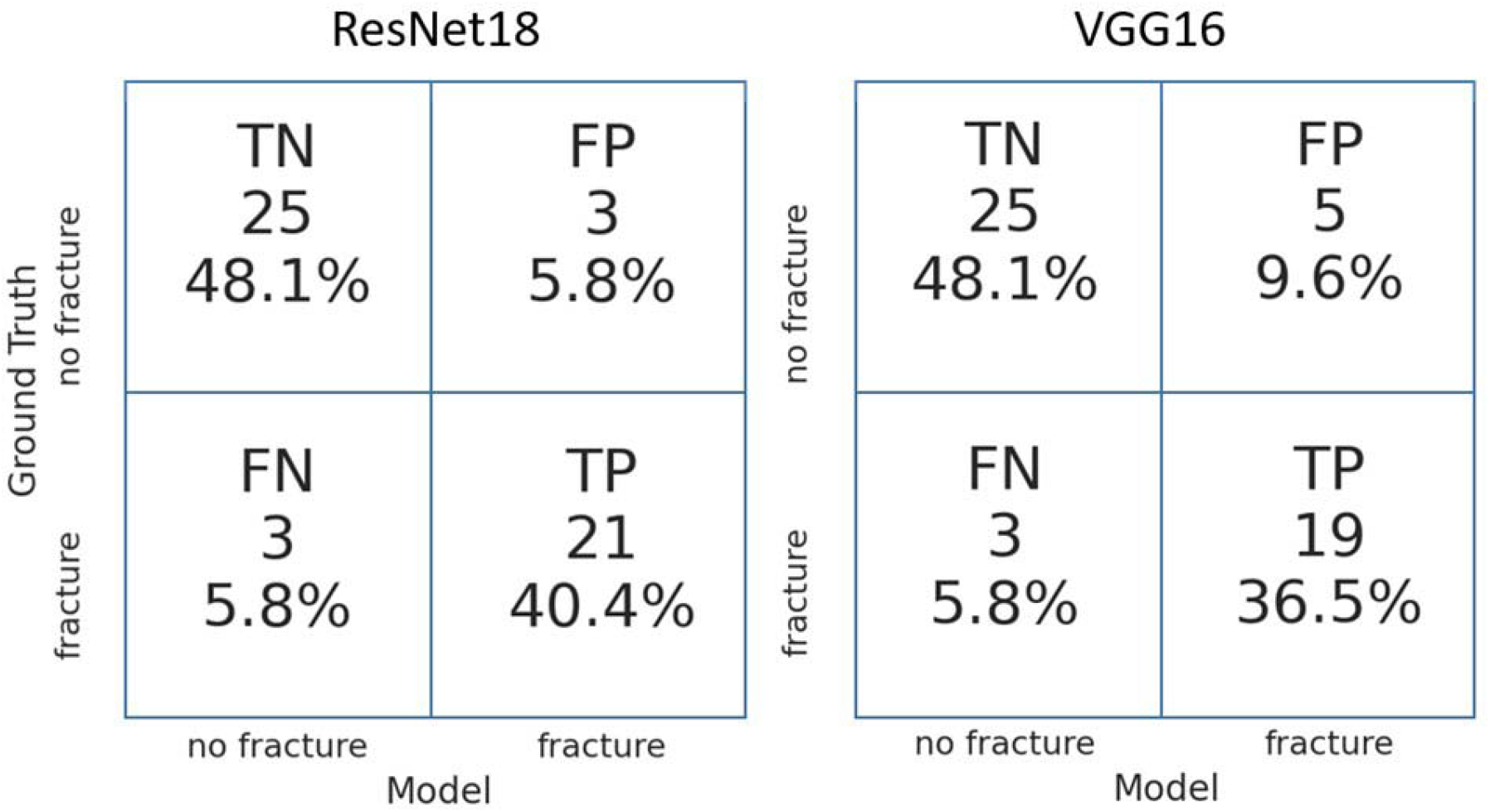
Confusion matrices obtained with the two DL architectures. The ResNet18 model made 6 misclassifications, whereas VGG16 made 8.TN: True negative; FN: false negative; TP: true positive; FP; false positive

In terms of area under the ROC **(Fig. 5)** the ResNet18 performed better than the VGG16, with 0.88 and 0.86 respectively for both classes.

**Figure 5:**
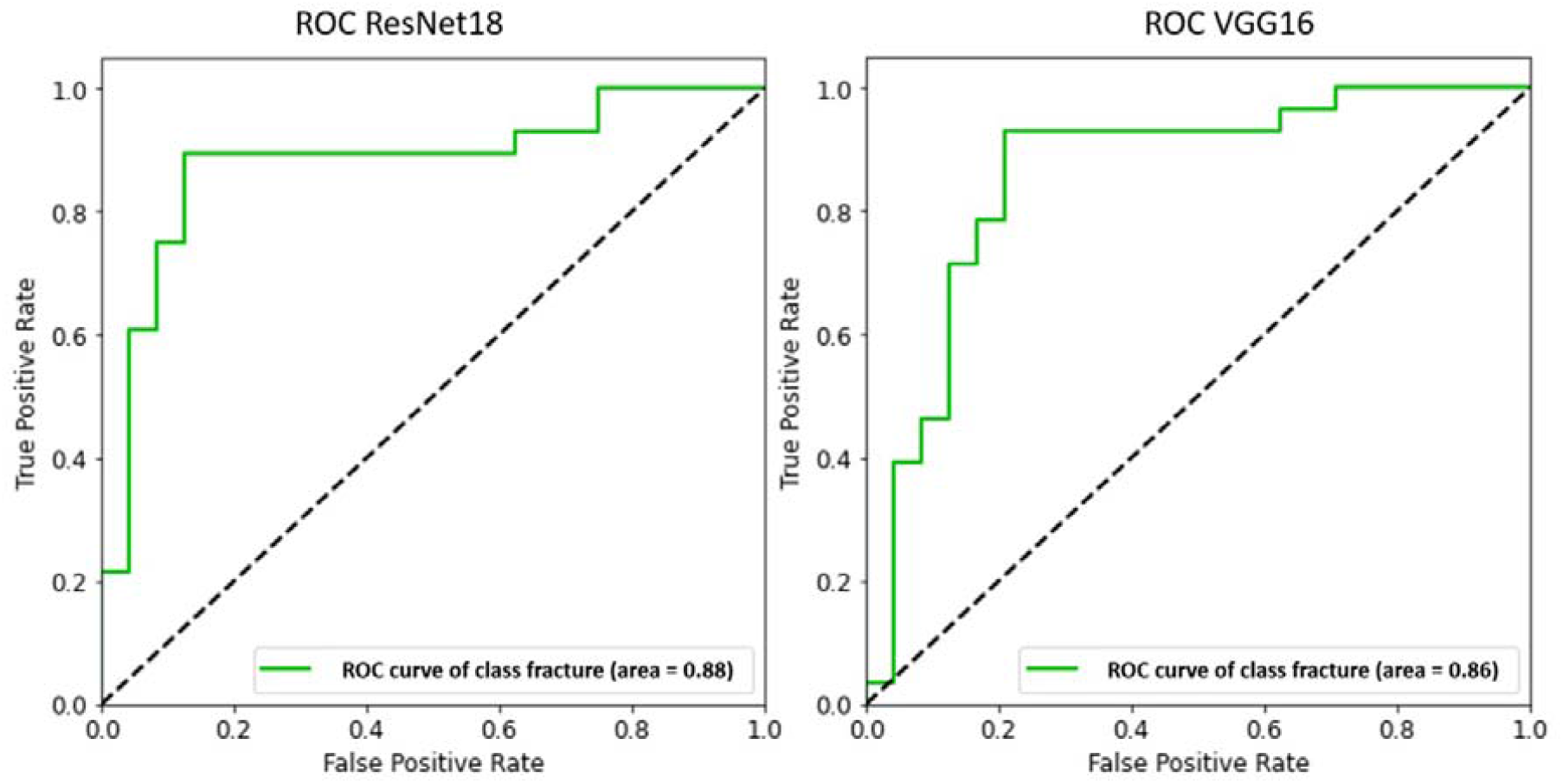
Comparison of the ROC curve obtained with ResNet18 and VGG16.

To ensure that the model’s prediction was based on the correct identification of the fracture zone, we conducted an Activation Map analysis. The resulting heatmaps depict which areas of the images led the model to classify the vertebra as ‘fracture’ or ‘no fracture’ **(Fig. 6)**. In 81% of the single vertebrae, the “warm zone” correlated to the fracture zone observed in the corresponding CT or MRI. Interestingly, in two occasions, the model’s prediction made the surgeons question the correctness of the ground truth. After verification via MRI and CT, the two images had to be reassigned to the opposite class. The model’s prediction effectively amended human errors made during the annotation process. Accounting for this reassignment, the number of false negatives would be reduced from 3 to 1, thus increasing the sensitivity from 89% to 96%.

**Figure 6:**
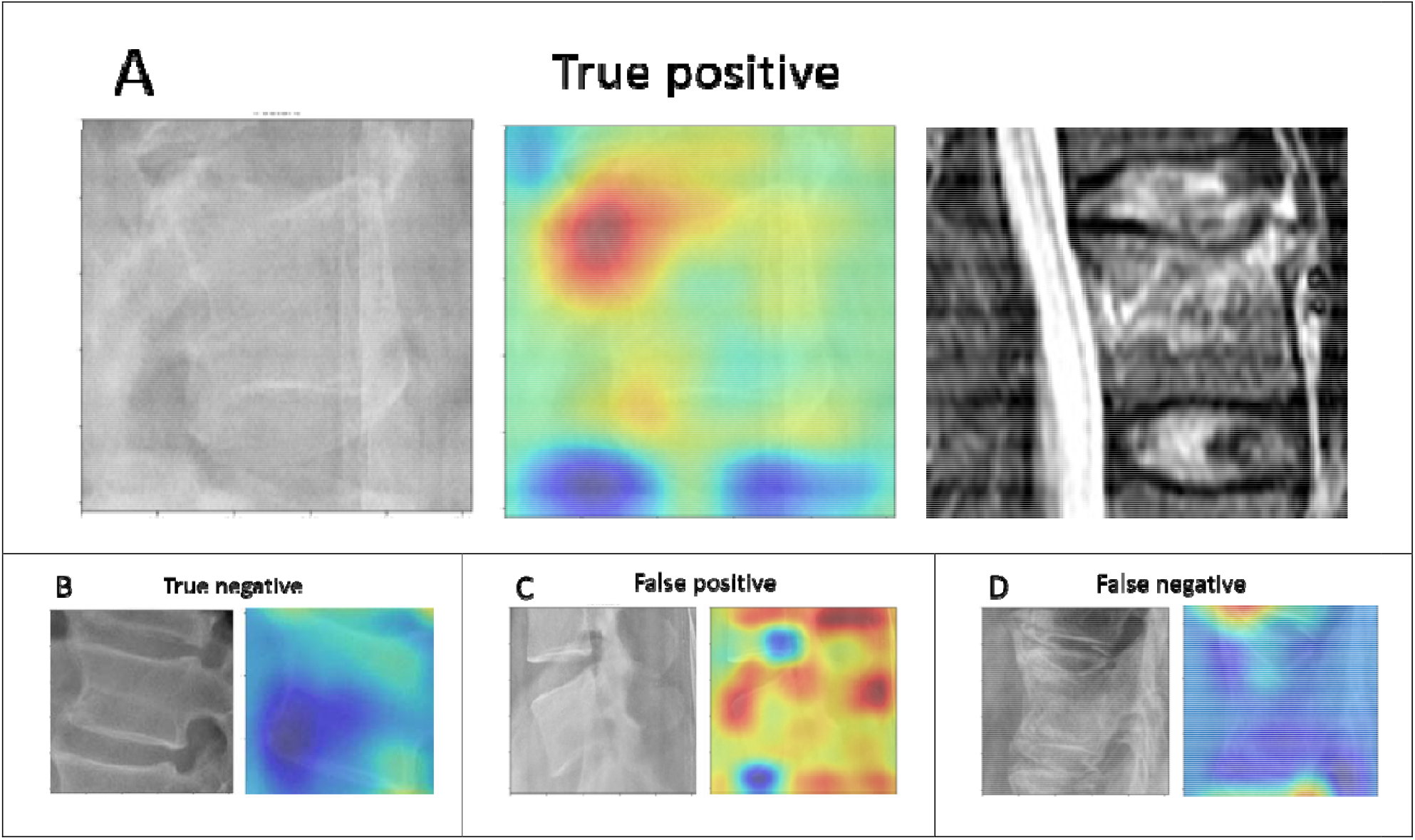
Heatmap analysis of the fracture zone. In Panel a, the heatmap correlates to the fracture zone identified in MRI. Panel b shows a true negative where the heatmap did not highlight a critical zone on the vertebra. Panel c incorrectly indicates presence of a fracture. Panel d was originally classified as ‘fracture’ and thus accounted for as a false negative, but it was then reclassified as ‘no-fracture’ after the evaluation of the heatmap.

## Discussion

This study demonstrated that AI-based techniques can detect vertebral fractures on radiographs with very high accuracy. Both models achieved similar sensitivity and specificity to that achieved by expert surgeons and radiologists (26,33–37) and the average sensitivity reported in a recent review.(38) ResNet18 showed better performance in identifying vertebral fractures compared to VGG16. Additionally, ResNet18 was less resource intensive in terms of memory used by the network parameters (43 MB vs 524 MB) and faster in the inference. To our knowledge, this is the first time that ResNet18 has been adapted for fracture identification purposes. Furthermore, ResNet18 predictions were based in most cases, on the regions of vertebrae corresponding to the fracture zone observed in the CT and MRI. To ensure the quality of the diagnostic trial, the reference standard was established only after confirmation on CT and/or MRI by three different experts, as suggested by a recent meta-analysis of the diagnostic accuracy of deep learning in orthopedic fractures (38) and the expert panel recommendations from the Radiological Society of North America. (39)

### Comparison of Model’s Performance

Only recently have AI-based models been used to attempt fracture detection on radiographs. Presumably the first proof-of-concept paper using CNNs for fracture identification was published by Olczak in 2017. They compared the fracture identification capability of 5 existing CNNs. Fracture presence was deduced by extracting a combination of expressions and keywords from the radiologist’s report, namely metadata. Contrasting to our results, VGG16 showed the best performance in their study, achieving 83% fracture identification accuracy. (33)

Other authors have also adapted CNNs to the problem of fracture detection, focusing exclusively on image interpretation, namely the information within the image file. Kim and MacKinnon used an adapted version of the Inception V3 model to identify distal radius fractures on sagittal radiographs, achieving an AUC of 0.954. (36) Although a significant limitation of this previous study was the exclusion of radiographs if the single lateral projection was inconclusive for the presence of fracture, their model analyzed the complete radiograph image instead of a cropped region of interest, as we and most other researchers have done. Chung et al used an adapted ResNet-152 on cropped anteroposterior radiographs of the shoulder to distinguish fractured from normal humeri, achieving an accuracy of 95%, AUC of 0.996, sensitivity of 99% and specificity of 97% in the optimal cutoff point. (40) Adams et al. used cropped radiographs of surgically confirmed femoral neck fractures to compare the performances between AlexNet and GoogLeNet. GoogLeNet outperformed AlexNet, achieving an overall accuracy of 90.6%. Given that the reference standard was established surgically, the probability of bias introduction into the model was cleverly minimized. (34) Similarly, we minimized annotation bias by training the model exclusively with radiographs where the presence of the fracture was confirmed via CT or MRI. Urakawa et al also evaluated cropped radiographs of femoral neck fractures and achieved an accuracy of 95.5%, AUC 0.984, sensitivity of 93.9% and specificity of 97.4% using an adapted version of VGG16. (37)

Recently, a model based on Visual Recognition V3 (IBM, Armonk, NY, USA) was used to identify vertebral fractures by Murata et al, achieving an accuracy, sensitivity, and specificity of 86.0%, 84.7%, and 87.3% respectively. (26) While their results are similar to ours, there are important methodological differences to consider. To avoid introduction of systematic errors while training the model, all the fractures included in our study were evaluated individually by expert spine surgeons before annotation, and then discussed in consensus meetings where discrepancy occurred. In contrast, each classifying surgeon in the study of Murata et al. seemingly evaluated a single subgroup of images. While our model was trained to identify anomalies in single vertebrae to eliminate confounding factors and ensure a future clinical applicability, as shown in the heatmap analyses **(Fig. 5)**, Murata’s group analyzed the entire radiograph. The exclusion of cases with multiple traumatic fractures impairs the application of their model in the clinical practice. However, the inclusion of anteroposterior radiographs approaches a regular clinical scenario where both projections would be evaluated. In addition to the use of a different model, these factors might have contributed to the marginally better performance achieved in our study.

### Heatmap Analysis

In 81% of the cases, our model’s prediction of ‘fracture’ or ‘no fracture’ was based on a precise identification of the anomalies in single vertebrae, confirmed by correlating the “warmer zones” with the findings in CT and/or MRI **(Fig. 6)**. Interestingly, two images which the model predicted as ‘no fracture’ by the model were originally classified as ‘fracture’ by the surgeons. A reassessment of the images supported the model’s prediction and increased the model’s sensitivity to 96%. Although this finding should be cautiously considered due to its exemplary nature, it illustrates the potential of AI to contribute to physicians’ decisions in the clinical workflow.

### Limitations

The present study has limitations. First, the dataset had a relatively small size. Traumatic vertebral fractures are commonly diagnosed based on CT or MRI only, obviating the need for radiographs in most cases. Although a larger database would arguably have enhanced the performance of the CNNs, we mitigated the impact of this limitation by performing aggressive image augmentation and taking advantage of models pre-trained on the ImageNet dataset. Since in the clinical workflow, a surgeon or physician would mostly rely on CT and MRI to confirm the presence of a fracture, a comparison with a model trained only with sagittal radiographs seemed unbalanced for this study’s purpose. A comparative evaluation of the performance of the classifier and that of surgeons who are naïve to the clinical images will be reported in a future study. Regarding the heatmaps, it should be noted that the activation maps do not necessarily show the fracture zone but rather the zones that are more important in determining the output of the classifier, which may not correspond to the fracture itself.

### Clinical Relevance of AI for automated traumatic lesion detection

Introducing systems of radiograph interpretation can reduce the frequency of misdiagnosis to below 0.3%.(41) Failures in fracture identification can be considerably reduced by implementing a second-stage verification algorithm at the end of the normal workflow to complement the interpretation of the physician. This way, introduction of bias or distractions would be avoided.

Contrary to common belief, computer-aided diagnostic tools are not necessarily aimed to replace expert human interpretation of medical imaging. In the authors view, the goal is minimization of Diagnostic Errors. Currently up to 30% vertebral fractures in radiographs are missed (1–3), resulting in either delayed or missed diagnosis. Both outcomes are qualified as Diagnostic Errors by the Institute of Medicine (42) and carry important legal and clinical implications:

- Legally, misdiagnoses are the most common source of malpractice claims or litigation.(43)
- Clinically, missed fractures in radiographs have consequences such as malunion with restricted range of motion, posttraumatic osteoarthritis, and joint collapse (44)

Physicians commonly tackle these implications by performing confirmative CT and/or MR studies, inevitably resulting in a delay in diagnosis, increase in costs and potentially also higher exposure to radiation. The delays can range from hours to months, resulting in poorer clinical outcomes. (10,11)

A commonly mentioned rebuttal for the implementation of AI based algorithms is the so called “black box” problem, where the clinician is blinded to the “reasoning” behind the model’s prediction. (45) Visualization techniques such as heatmaps could improve the acceptance of fracture detection systems in the clinical practice.

## Conclusion

This study found that our AI model can accurately identify anomalies suggestive of thoracolumbar vertebral fractures in sagittal radiographs. Specifically, an adapted version based on ResNet18 achieved a similar performance compared to other models, and those reported of expert surgeons and radiologists. Additionally, it also highlighted a human error made during the annotation process. Applying this AI model to minimize diagnostic errors in fracture detection in sagittal radiographs of the TL vertebra seems plausible.

## Data Availability

The data that support the findings of this study are available from the corresponding author, GSR, upon reasonable request.

